# Psychological distress and compliance with sanitary measures during the Covid-19 pandemic: effect modification by participants’ gender and socioeconomic characteristics, an IPD meta-analysis

**DOI:** 10.1101/2025.01.06.24319678

**Authors:** Irwin Hecker, Solène Wallez, Honor Scarlett, José Luis Ayuso-Mateos, Richard Bryant, Giulia Caggiu, Claudia Conflitti, Katalin Gémes, Josep Maria Haro, Vincent Lorant, Roberto Mediavilla, Ellenor Mittendorfer-Rutz, Anna Monistrol-Mula, Matteo Monzio Compagnoni, Papoula Petri-Romão, Irene Pinucci, Marit Sijbrandij, Jutta Stoffers-Winterling, Henrik Walter, Murielle Mary-Krause, Maria Melchior

## Abstract

**Background:** This study aims to understand how psychological distress is related to compliance with COVID-19 sanitary measures. In addition, we explored whether gender and socioeconomic status (i.e educational level and employment status) can modify this relationship.

**Methods:** Data from four European cohort studies (n=13,635), were analysed using an Individual Participant Data (IPD) meta-analytic approach. Mixed effect models were employed to examine associations between mental health difficulties and compliance with sanitary measures, as well as effect modification by socioeconomic status. Statistical models were additionally stratified by gender.

**Results:** We found a statistically significant positive association between mental health difficulties and compliance with sanitary measures in women, while amongst men the statistically significant association observed was negative. Moreover, there was a statistically significant interaction between participants’ educational level and mental health difficulties amongst men only, indicating even lower compliance levels with COVID-19 sanitary measures amongst individuals with only primary schooling.

**Conclusion:** The association between psychological distress and compliance with sanitary measures is complex - positive in women, negative in male. Men experiencing mental health difficulties, especially those with lower educational attainment, exhibit low levels of compliance with sanitary measures.

## Introduction

Echoing patterns seen during prior epidemics [1], the COVID-19 pandemic showed widespread adverse impacts on global mental health globally [2–6]. The implementation of sanitary measures played a crucial role in mitigating the pandemic [7]. People’s compliance with sanitary measures is also very important as it impacts their effectiveness regarding the epidemiological impact [8].

The relationship between psychological distress and compliance with sanitary is uncertain from a theoretical perspective, with very few studies having compliance as the outcome. Two possibilities, positive and negative relationships, might arise. It could be positive, assuming increased compliance due to anxiety as it has been shown that people with mental health problems had lesser tendency to stop social distancing even as the pandemic subsided. They may surpass the recommended precautions and take them to an extreme [9]. Or negative, as compliance with sanitary measures might be a challenge too difficult to face for people with psychological distress that might already face gendered socioeconomic difficulties. Indeed, evidence shows that mental health follows a socioeconomic gradient [10–12]. Having a lower socioeconomic position affects mental health through both material and psychosocial factors [12]. This was also the case during the COVID-19 pandemic, where people experiencing financial difficulties or unemployment were at higher risk of experiencing symptoms of anxiety/depression compared to those who did not [13,14]. Gender inequalities were also aggravated within the context of the pandemic. Women were already more likely to endure psychological distress, such as anxiety and depression [15], before the pandemic, which further aggravated pre-existing gender inequalities (e.g., unpaid work disparities, disproportionate caregiving burden) compared to men [16,17]. For instance, on the one hand, employed women were particularly at risk for their mental health given that they often carried the burden of domestic responsibilities, such as caring for children and the elderly, as well as changes in work-life balance [16,17]. On the other hand, women aged 35-39 years experienced higher levels of pandemic-induced unemployment, with less educated women with young children particularly at risk [18].

Moreover, sanitary measures (e.g., lockdowns and business closures, remote work mandates, social distancing regulations, travel or healthcare access restrictions, school and university closures, and border closures)[19] also have uneven economic and social impacts on people [20]. Whilst these measures were proven to mitigate COVID-19 outcomes, they exacerbated socioeconomic inequalities [21,22] due to factors such as heterogeneity in one’s ability to work remotely or the probability of COVID-19 infection within the household [23]. Individuals with no formal education and those with only primary education appear less aware of available mental health care services [24]. Being employed [25] and having a low educational level [25–28] also associate with non-compliance with sanitary measures because of other priorities such as securing means of subsistence [25]. Then people may be exposed to unsafe work conditions or unstable income resources [25,29–31]. Regarding gender differences, behaviours also differ. More specifically women tend to be more compliant with preventive measures (e.g., reduced mobility or wearing masks) than men [32].

Based on this context, we anticipated that compliance with sanitary measures during the COVID-19 pandemic would be a challenge for certain populations. Drawing from the observed gender differences in compliance with sanitary measures during the COVID-19 period, and their association with psychological distress, we hypothesised that the association between compliance with sanitary measures and psychological distress would be different across genders. Also, educational level and employment status might have played a distinct role in shaping these dynamics for both men and women.

## Methods

### Study design and population

To investigate this, we combined data from multiple datasets collected during the COVID-19 pandemic into an Individual Participant Data (IPD) dataset. In our research, we employed IPD meta-analysis [33], utilising datasets exclusively sourced from partners within the RESPOND project. We carefully curated target variables and harmonised these partner datasets. Rigorous quality checks were conducted to ensure the reliability and integrity of the data. This approach enhances the depth and reliability of our findings, enabling robust conclusions to be drawn from the combined datasets contributed by RESPOND project partners.

The population for this study relied on data collected from four observational cohort studies: (i) the TEMPO (Trajectoires ÉpidéMiologiques en Population)[34], (ii) MINDCOVID [35], (iii) COVID and I [36], and (iv) the COVID-19 Mental Health Survey (COMET)[37].

The TEMPO cohort began in 2009 with the aim of better understanding mental health patterns and addictive behaviours. Starting from 2020, TEMPO participants were contacted to collect data regarding their health status during the pandemic. Nine waves of data were collected using self-administered questionnaires from March 24, 2020 (one week after the beginning of the first lockdown), to the end of July 2021. All COVID-19 data waves have been included in this study.

A baseline survey of a cohort of general population adults was conducted as part of the MINDCOVID project. The target population consisted of non-institutionalised Spanish adults (i.e. aged 18 years or older) without Spanish language barriers. Professional interviewers carried out computer-assisted telephone interviews (1–30 June 2020) in a sample drawn using dual-frame random digit dialling. Only the first and second waves of data collection were included in this study as questions related to compliance were asked at these waves only.

An online web survey, named COVID and I, was distributed across Belgium through social media and national news outlets during the beginning of the first wave of the COVID-19 pandemic in 2020. The Belgian government implemented their first restriction measures on March 13th, the survey was launched on March 20th, two days after the start of the lockdown. The survey was aimed at the general population and was available in English, French, and Dutch. Only the fourth wave of data collection was included in this study as questions related to compliance were asked at that wave only.

The COMET study is an international, online longitudinal survey aimed at evaluating the course of mental health symptoms during the COVID-19 pandemic, as well as identifying individuals at greater or reduced risk of mental illness. The COMET sample includes participants from 14 countries (The Netherlands, Italy, Switzerland, Turkey, Spain, Germany, France, the United Kingdom, Sweden, South Africa, Indonesia, China, Australia and the United States). Participants were recruited starting in May 2020 through a snowball sampling strategy using university mailing lists and different social media platforms. Only the fourth and fifth waves of data collection were included in this study as questions related to compliance were asked at these waves only.

Database data collection is detailed in Supplementary Figure 1 and population selection in Supplementary Figure 2. Other RESPOND databases (EDAD CON SALUD, HEROES, LORA, MARP, DYNACORE-L) were not included because they had no information regarding compliance (EDAD CON SALUD, HEROES, LORA, MARP) or mental health (DYNACORE-L).

### Ethics

All studies have been approved by the appropriate ethics committee and have therefore been performed in accordance with the ethical standards laid down in the 1964 Declaration of Helsinki and its later amendments.

The COMET study was approved by the ethical review board of the Faculty of Behavioral and Movement Sciences of the Vrije Universiteit Amsterdam (VCWE-2020-077). Personal data is protected according to EU and national laws.

Covid and I’s ethical review and approval was not required as it is a population-based, online survey without the collection of personal data, which is in accordance with the local legislation and institutional requirements. Participants were provided with the legal information relating to consent, and online informed consent was obtained from all participants.

The MINDCOVID study protocol was approved by the IRB Parc de Salut Mar (2020/9203/I) and by the corresponding IRBs of all the participating centres. The study is registered at ClinicalTrials.gov (https://clinicaltrials.gov/ct2/show/NCT04556565).

The TEMPO cohort received approval of bodies supervising ethical data collection in France, the Advisory Committee on the Treatment of Information for Health Research (Comité consultatif sur le traitement de l’information en matière de recherche dans le domaine de la santé, CCTIRS) and the French regulatory data protection authority (Commission Nationale de l’Informatique et des Libertés, CNIL, n⍰ 908163).

All data included in the present analyses were fully anonymized.

## Measures

### Outcome

Compliance with sanitary measures was measured using different using self report items (handwashing, social distancing, physical contact, wearing a mask, lockdown, working from home, limiting small and/or large gatherings, curfew, hosting people at home, quarantine, and taking extra precautions with at risk people). These items were harmonised by summing them (Supplementary Table 1) and calculating z-scores within each sample.

### Exposure

Psychological distress were measured using different scales according to each considered cohort study (COMET: Patient Health Questionnaire Anxiety and Depression Scale (PHQ-ADS)[38]; COVID and I: General Health Questionnaire-12 (GHQ-12)[39]; MINDCOVID: Patient Health Questionnaire-8 and the Anxiety and Depression Scale (PHQ8-ADS)[40]; TEMPO: Adult Self-Report (ASR))[41] and harmonised using a standard procedure: first, for each scale, the different items (Supplementary Table 2) were summed up, and after the corresponding z-scores were calculated. In this way, the individual values of each single scale across the four studies were transformed into a measure of the same order of magnitude (numeric, with specific minimum and maximum), making them comparable. For each individual, regardless of the cohort he or she belongs to, the corresponding z-score obtained from the cohort-specific evaluation scale was considered as the exposure.

### Covariates

Sociodemographic characteristics were collected across the four studies, encoded consistently using the same names, values, and formats and included: gender (“Female”; “Male”), age, education (“Tertiary”; “Primary”; “Secondary”), employment status (“In employment”; “Unemployed”), and number of children (“No children”; “One”; “Two or three”; “Four or more”).

All variables accounted for in this study had previously been reported as factors associated with compliance with sanitary measures: female gender [28,42–46], old or young age [26,43,44,46–50], education [25–28], employment status [51] and number of children [52].

A “stringency” variable was also included, based on the Stringency Index [53]. The Stringency Index incorporates nine metrics: school, workplace, or public transport closures, the cancellation of public events, restrictions on public gatherings, restrictions on internal movements, stay-at-home requirements, public information campaigns, and international travel controls. The score corresponds to the mean score of these nine metrics, with each metric ranging from 0 to 100. A higher number on the Stringency Index represents a stricter response from the country where the participant resides, with 100 indicating the strictest possible measures.

### Statistical analysis

To test associations between participants’ psychological distress, socioeconomic status (i.e educational level and employment status) and compliance with the sanitary measures implemented during the COVID-19 pandemic, individual participant data from relevant studies was merged and analysed. Due to the longitudinal nature of the data, mixed effect models were used to calculate adjusted odds ratios (aOR), and the corresponding 95% confidence intervals (CI). Multivariate mixed effect models were adjusted for the above-listed covariates (please see the “Covariates” subsection). Analyses were conducted individually for each database, as well as for the merged dataset. Data were stratified by gender to consider gender-specific patterns related to the outcome, exposure, and socioeconomic status. Additionally, interactions between socioeconomic status and exposure were explored. P-values of less than 0.05 were considered to be statistically significant. Collinearity of model variables was explored and measured using Variance Inflation Factor (VIF) derivatives, namely generalised VIF (GVIF) and GVIF^(1/(2*Df))[54–57]. After removing incomplete cases for both the outcome and exposure, 19,143 longitudinal observations (from 13,635 participants) had data sufficient for analyses. Incomplete data on covariates, with an average of 5% missing data, were imputed using Multiple Imputations by Chained Equations (MICE) with Fully Conditional Specification (FCS), based on five multiple imputations [58,59]. All statistical analyses were performed using R version 4.2.3 and Rstudio version 2023.6.1.524 [60].

## Results

The four included cohorts showed many similarities, including a higher proportion of women (68.9%), and higher rates of both employment (83.5%) and tertiary education (83.7%). The mean age in each cohort ranged from 40 to 46 years. Among TEMPO participants, there was a high proportion of individuals with two or three children (57.9%), while most participants of COMET, COVID and I and MINDCOVID, had no children (respectively 56.6%, 40.9%, 54.8%). The mean stringency score ranged from 55 for COMET to 81 for TEMPO (Table 1).

Before stratifying by gender, the association between psychological distress and compliance with sanitary measures was statistically non-significant (aOR 1.01, 95%CI: 0.99 - 1.02).

When stratifying data by gender and adjusting for covariates, the association between psychological distress and compliance with sanitary measures was negative in men (aOR: 0.93, 95%CI: 0.90 - 0.97) and positive in women (aOR: 1.03, 95%CI: 1.01 - 1.06) (Figure 1).

Additionally, we found a statistically significant interaction between psychological distress and primary education among men (aOR: 0.82, 95%CI: 0.68 - 0.99) (Figure 2), but not for the other categories of education and employment status (Figure 3).

## Discussion

### Main findings of this study

Data collected among the 13,635 participants of the COMET, COVID and I, MINDCOVID and TEMPO cohorts from March 2020 to August 2022, revealed an association between psychological distress and compliance with sanitary measures during the COVID-19 period when stratified by gender. These gender differences revealed that women experiencing psychological distress showed higher compliance with sanitary measures, whereas the opposite relationship was observed amongst men. Additionally, an interaction between psychological distress and educational level among men in the association with compliance with sanitary measures was observed, indicating an even lower level of compliance for this group.

### What is already known on this topic

The influence of both gender and socioeconomic characteristics on the association between psychological distress and compliance with sanitary measures is not yet well documented. Still, previous studies can explain how psychological distress has different effects on compliance across genders. Indeed, gender disparities in risk-taking behaviours [61] and health-related decision-making patterns during the COVID-19 pandemic have been documented [62]. Globally, men are more likely to engage in risky behaviours, and less inclined to seek preventive medical care, or support for health issues [63–65]. During the pandemic, men perceived the consequences of COVID-19 to be less severe compared to women, despite objective evidence suggesting otherwise. Traditional masculinity norms appear to moderate this perception, which, in turn, negatively affects adherence to protective measures [66].

### What this study adds

Our study has a number of strengths worth highlighting. A significant strength of our study lies in the IPD meta-analytical approach, with which we were able to include four large cohort studies. IPD meta-analyses are recognized for providing a more comprehensive assessment of pooled data when compared to aggregate data analyses [67]. This methodology also allowed us to extract and analyse raw data from each individual study, including diverse spatial and temporal contexts throughout the COVID-19 pandemic, and thus enhance the precision and robustness of our findings by taking into account various contexts of data collection.

An interaction between psychological distress and educational level in terms of compliance with COVID-19 sanitary measures was observed among men in our study. This suggests an even lower level of compliance within this group. It is known that bi-directional effects between academic achievement and social withdrawal exist in boys thereby increasing the risk of psychosocial maladjustment, depression, loneliness and anxiety [68]. This may partially explain the subsequent difficulty in complying with measures later in life for this population of low educated men.

Containment policies have resulted in a reduction of the impact of COVID-19 [69,70]. Sanitary measures have been proven to contain the spread of the virus [70,71], showing a linear, inverse relationship between the incidence of COVID-19 and degree of observed prevention measures [72]. During a health crisis, awareness of groups with low compliance levels can significantly help in proactively intervening, especially considering that shorter lockdown periods can be compensated for by high adherence to interventions, leading to similar epidemiological impacts [8]. It is, then, indeed important to educate the public about the negative consequences of the virus and the effectiveness of sanitary measures [73]. Regarding mental health, recommendations should aim to reduce mental health inequalities between vulnerable groups and the general population using measures targeted and adapted to specific contexts [74].

As it has already been recommended, a gender-specific response to a new health risk emphasises the need for targeted public health messaging [32]. In a recent scoping review [62], it was revealed that people’s perceptions of COVID-19 health information and recommendations, as well as their decision-making regarding health matters, are significantly influenced by their level of education and health literacy. This is particularly important since the effectiveness of COVID-19 containment measures relies on widespread public understanding and support. Targeting low educated men with tailored mental health interventions would not only help tackle psychological distress related to the pandemic but also promote compliance with sanitary measures, thus reducing the risk of infection [75]. Mental health interventions for vulnerable groups are currently being tested within the context of the COVID-19 pandemic and its aftermath [76–79].

### Limitations of this study

We need to acknowledge some limitations to our study. As the data has been self-reported, it may contain biases stemming from social desirability and memory recall issues [80]. Some key variables, such as participants’ income [27,44,47,50,81], chronic illness [82], or COVID-19-related worries [26] were excluded due to their heterogeneity across all included studies. However, this selective inclusion was a deliberate choice aimed at maintaining methodological consistency across studies and ensuring rigorous analysis. By focusing on shared variables, we enhanced the internal validity of our study, thereby providing a more reliable synthesis of the available evidence. Both outcome and exposure data were derived from distinct validated scales and questions within each survey, which were subsequently pooled post-collection using methods distinct from the standard approaches for the respective scales. This set of data is not comprehensive as it was not identified and selected systematically; rather, it was included from the datasets provided by the partners of the RESPOND project. Nevertheless, this approach allowed a harmonised comparison of different datasets.

## Conclusion

Our study underscores the need for targeted interventions related to compliance with preventive measures among persons experiencing psychological distress, particularly during health crises like the COVID-19 pandemic. Our findings highlight the importance of tailoring messaging and strategies to address the unique challenges faced by different populations. Targeting specific groups with lower rates of compliance through tailored messaging is essential for effective management of health crises, such as the COVID-19 pandemic. Men experiencing psychological distress, especially those with lower educational attainment, had a lower compliance with sanitary measures. This involves a targeted approach towards men experiencing psychological distress, especially those with lower educational attainment. By promoting the well-being of all individuals, taking into account mental health, gender, and socioeconomic factors, we may better prepare for future health crises.

## Data Availability

Data cannot be shared publicly because of data protection regulations. Data are available from the Pierre Louis Institute for Epidemiology and Public Health (IPLESP) for researchers who meet the criteria for access to confidential data.

## Funding

COMET received no external funding.

Covid and I is supported by a grant from the King Baudouin Foundation, Grant No.: 2020-J1812640-216406.

MINDCOVID is supported by Instituto de Salud Carlos III (Ministerio de Ciencia e Innovación)/FEDER (COV20/00711); ISCIII (Sara Borrell, CD18/00049) (PM); FPU (FPU 15/05728)); ISCIII (PFIS, FI18/00012); Generalitat de Catalunya (2017SGR452).

The TEMPO cohort is supported by the French National Research Agency (ANR), the French Institute for Public Health Research-IReSP (TGIR Cohortes), the French Inter-departmental Mission for the Fight against Drugs and Drug Addiction (MILDeCA), the French Institute of Cancer (INCa) and the Pfizer Foundation.

This study is supported by the European Union’s Horizon 2020 research and innovation program RESPOND (The RESPOND project is funded under Horizon 2020 – the Framework Programme for Research and Innovation (2014– 2020). The content of this article reflects only the authors’ views and the European Community is not liable for any use that may be made of the information contained therein).

## Ethics declarations

### Conflict of interest

The authors declare that they have no conflict of interest.

## Notes

### Competing Interest Statement

The authors have declared no competing interest.

### Funding Statement

Yes

