## Supplementary material for "Psychological distress and compliance with sanitary measures during the Covid-19 pandemic: effect modification by participants’ gender and socioeconomic characteristics, an IPD meta-analysis": Tables

Table 1: Characteristics of COMET, COVID and I, Mind COVID, and TEMPO cohort studies, March 2020 - August 2022, n=13,635.

| **Characteristics** | **COMET** (25.03%)*^1^* | **COVID and I** (30.97%)*^1^* | **MINDCOVID** (23.58%)*^1^* | **TEMPO** (20.42%)*^1^* | **All datasets***^1^* |
| --- | --- | --- | --- | --- | --- |
| N | 3835 | 5935 | 2974 | 891 | 13,635 |
| Gender |  |  |  |  |  |
| Female | 79.6% | 74.7% | 53.1% | 65.3% | 68.9% |
| Male | 20.4% | 25.3% | 46.9% | 34.7% | 31.1% |
| Age in years | 42 (16) | 44 (12) | 46 (14) | 40 (4) | 43 (12) |
| Education |  |  |  |  |  |
| Tertiary | 75.0% | 89.3% | 80.4% | 89.5% | 83.7% |
| Primary | 5.6% | 0.3% | 4.0% | 0.0% | 2.4% |
| Secondary | 19.4% | 10.3% | 15.7% | 10.5% | 13.9% |
| Employment Status |  |  |  |  |  |
| In employment | 85.3% | 85.6% | 75.5% | 87.6% | 83.5% |
| Unemployed | 14.7% | 14.4% | 24.5% | 12.4% | 16.5% |
| Number of Children |  |  |  |  |  |
| No children | 56.6% | 40.9% | 54.8% | 24.0% | 44.6% |
| One | 14.2% | 20.3% | 18.2% | 16.3% | 17.5% |
| Two or three | 26.5% | 35.6% | 26.1% | 57.9% | 35.6% |
| Four or more | 2.7% | 3.2% | 0.8% | 1.8% | 2.2% |
| Stringency | 55 (24) | 62 (2) | 60 (10) | 81 (11) | 64 (16) |
| *^1^%; Mean (SD).* | | | | | |

Figure 1: Association between mental health and compliance with sanitary measures in COMET, COVID and I, Mind COVID, TEMPO studies, and all populations, March 2020 - August 2022, n=13,635 (multivariate mixed models, adjusted odds-ratios (aOR), 95% confidence interval (CI)).

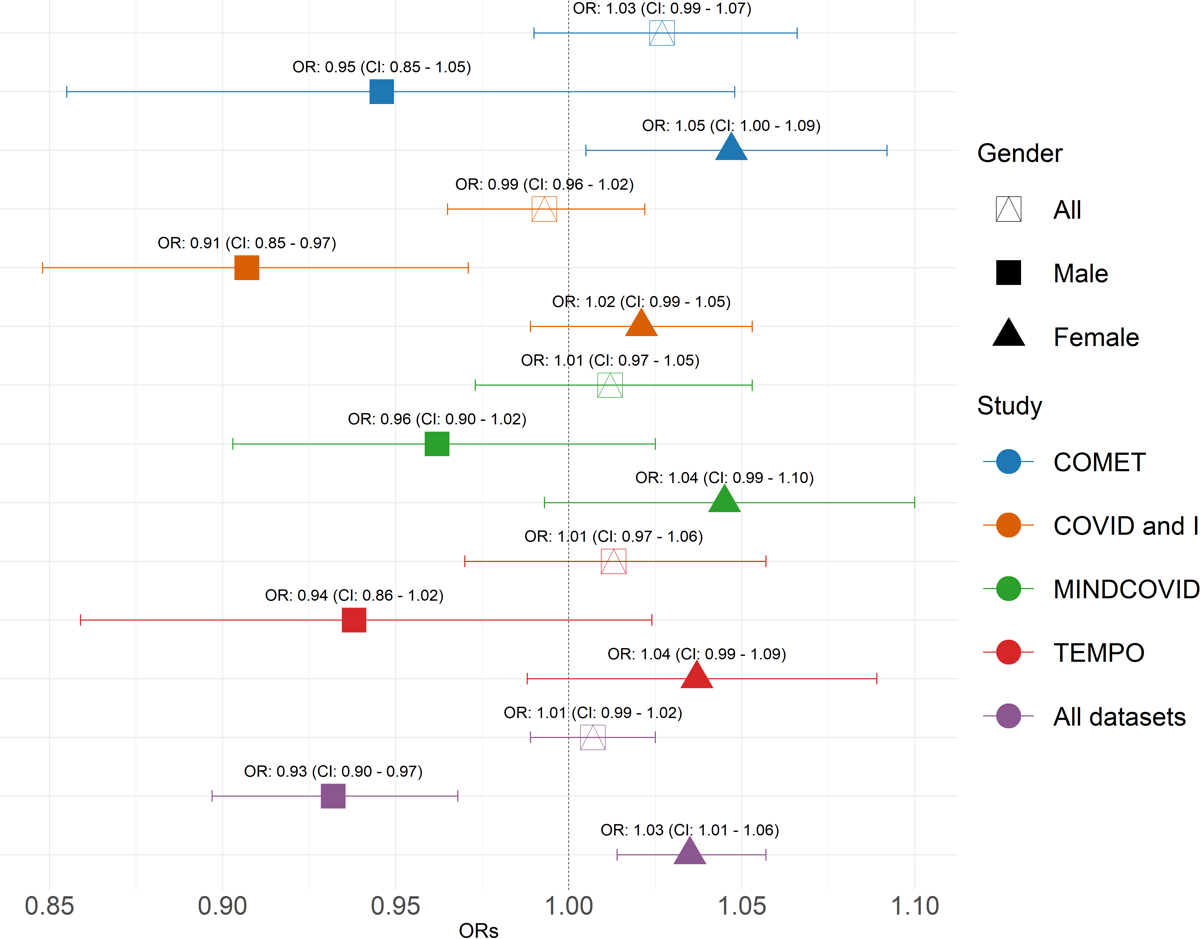

*All models adjusted for gender (except when stratified), age, education, employment status, number of children, and stringency. Both x and y axes show z scores.*

Figure 2: Interaction between participants’ educational level and mental health difficulties in relation to compliance with COVID-19 sanitary measures (COMET, COVID and I, Mind COVID, TEMPO studies), March 2020 - August 2022, n=13,635 (multivariate mixed models).

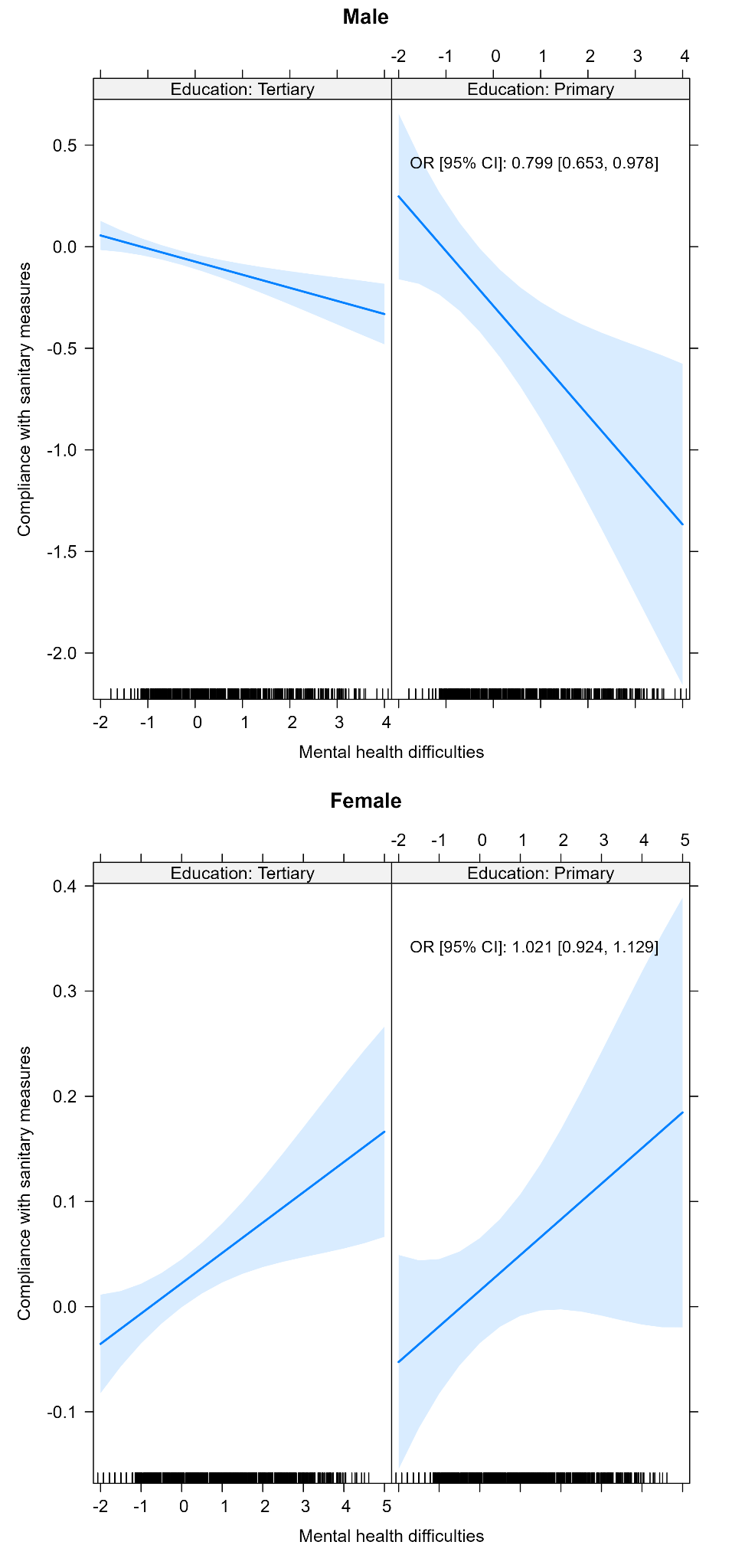

*All models adjusted for age, education, employment status, number of children, and stringency. Both x and y axes show z scores.*

Figure 3: Interaction between participants’ employment status and mental health difficulties in relation to compliance with COVID-19 sanitary measures (COMET, COVID and I, Mind COVID, TEMPO studies), March 2020 - August 2022, n=13,635 (multivariate mixed models).

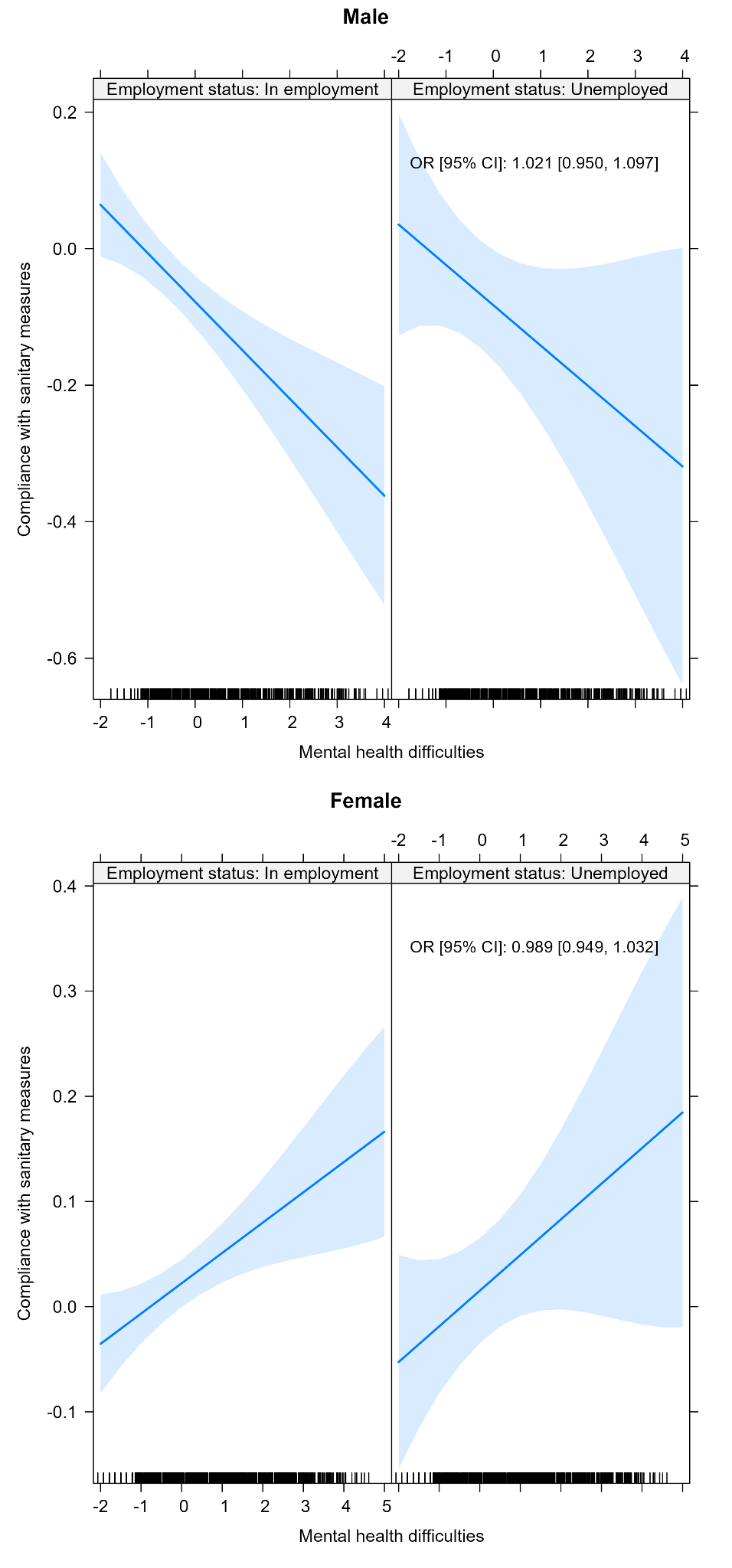

*All models adjusted for age, education, employment status, number of children, and stringency. Both x and y axes show z scores.*

Supplementary Figure 1: Data collected in the COMET, COVID and I, Mind COVID and TEMPO studies, March 2020 - August 2022, n=13,635.

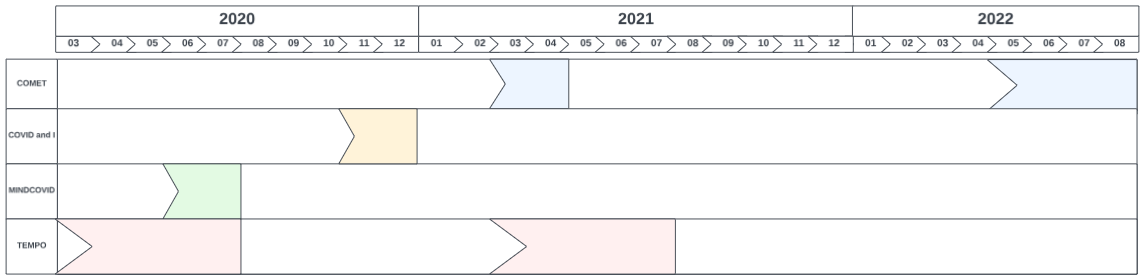

Supplementary Table 1: Items measuring compliance with sanitary measures against COVID-19 available in COMET, COVID and I, Mind COVID and TEMPO studies, March 2020 - August 2022, n=13,635.

| Study Name | Description | Question | Categories |
| --- | --- | --- | --- |
| COMET | Not being allowed to go outdoors, only with permission | How often did you adhere to the following regulations during the past week? | 1=Not at all  2=Most of the time  3=All of the time |
|  | Working from home |  |  |
|  | Restrictions related to small gatherings |  |  |
|  | Restrictions related to large gatherings |  |  |
|  | Wearing face masks in specific situations |  |  |
|  | Keeping a safe distance from people |  |  |
|  | Curfew |  |  |
|  | Being allowed to host only a limited number of people at home |  |  |
| COVID AND I | Maintain at least 1.5m distance from other people | To what extent have you complied with the following measures since their introduction? | 1=Strictly  2=Moderately  3=Scarcely  4=Not applicable |
|  | In case of illness: stay at home, do not go out shopping and/or receive visitors |  |  |
|  | Respect the “social bubble” (number of people allowed to meet with either at home or outside) |  |  |
|  | Take extra precautions with at-risk people |  |  |
|  | Cover your mouth and nose wherever it is compulsory |  |  |
|  | Cover your mouth and nose whenever at least 1.5 m distance from other people cannot be guaranteed |  |  |
| TEMPO Q1-Q7 | Increasing the frequency of handwashing (with soap or hand sanitiser) | What precautions do you take to avoid getting infected with Covid-19? Please tick all the boxes that apply to you. | 0 = Not ticked  1 = Ticked |
|  | Maintaining distance when passing by a stranger |  |  |
|  | Avoiding physical contact (including with family and friends) |  |  |
|  | Wearing latex gloves |  |  |
|  | Wearing a mask |  |  |
|  | Sneezing / coughing into your elbow |  |  |
|  | Avoiding public transport |  |  |
|  | Avoiding recreational and/or business trips |  |  |
| TEMPO Q8-Q9 | In public transport | In which situation do you respect shielding measures? Please tick every box that corresponds to your situation / Wearing a mask: |  |
|  | In the street |  |  |
|  | In shops |  |  |
|  | At work |  |  |
|  | When meeting friends |  |  |
|  | When meeting family |  |  |
|  | When meeting people at risk |  |  |
|  | In public transport | In which situation do you respect shielding measures? Please tick every box that corresponds to your situation / Handwashing: |  |
|  | In the street |  |  |
|  | In shops |  |  |
|  | At work |  |  |
|  | When meeting friends |  |  |
|  | When meeting family |  |  |
|  | When meeting people at risk |  |  |
|  | In public transport | In which situation do you respect shielding measures? Please tick every box that corresponds to your situation / Keeping a 1-metre distance: |  |
|  | In the street |  |  |
|  | In shops |  |  |
|  | At work |  |  |
|  | When meeting friends |  |  |
|  | When meeting family |  |  |
|  | When meeting people at risk |  |  |
|  | In public transport | In which situation do you respect shielding measures? Please tick every box that corresponds to your situation / No physical contact (hugging, kissing): |  |
|  | In the street |  |  |
|  | In shops |  |  |
|  | At work |  |  |
|  | When meeting friends |  |  |
|  | When meeting family |  |  |
|  | When meeting people at risk |  |  |
| MIND COVID | In the last 30 days, how do you rate your compliance with the restrictions imposed by the government? | | 1 = very low  2 = below average  3 = average  4 = above average  5 = very high |

Supplementary Table 2: Measures of mental health available in the COMET, COVID and I, Mind COVID and TEMPO studies, March 2020 - August 2022, n=13,635.

| **Study Name** | **Scale** | **Question** | **Description** | **Categories** |
| --- | --- | --- | --- | --- |
| COMET | PHQ9-ADS  (PHQ-9 and GAD-7) | Over the past 2 weeks, how often have you been bothered by any of the following problems? | Little interest or pleasure in doing things? | 0 = Not at all  1 = Several days  2 = More than half the days  3 = Nearly every day |
|  |  |  | Feeling down, depressed, or hopeless? |  |
|  |  |  | Trouble falling or staying asleep, or sleeping too much? |  |
|  |  |  | Feeling tired or having little energy? |  |
|  |  |  | Poor appetite or overeating? |  |
|  |  |  | Feeling bad about yourself—or that you are a failure or have let yourself or your family down? |  |
|  |  |  | Trouble concentrating on things, such as reading the newspaper or watching television? |  |
|  |  |  | Moving or speaking so slowly that other people could have noticed? Or the opposite—being so fidgety or restless that you have been moving around a lot more than usual? |  |
|  |  |  | Thoughts that you would be better off dead or of hurting yourself in some way? |  |
|  |  |  | Feeling nervous, anxious, or on edge |  |
|  |  |  | Not being able to stop or control worrying |  |
|  |  |  | Worrying too much about different things |  |
|  |  |  | Trouble relaxing |  |
|  |  |  | Being so restless that it is hard to sit still |  |
|  |  |  | Becoming easily annoyed or irritable |  |
|  |  |  | Feeling afraid, as if something awful might happen |  |
| COVID AND I | GHQ-12 | Have you recently... | ...been able to concentrate on whatever you're doing? | The positive items were corrected from 0 (always) to 3 (never) and the negative ones were corrected from 0 (always) to 3 (never). |
|  |  |  | ...lost much sleep over worry? |  |
|  |  |  | ...felt that you are playing a useful part in things? |  |
|  |  |  | ...felt capable of making decisions about things? |  |
|  |  |  | ...felt constantly under strain? |  |
|  |  |  | ...felt you couldn't overcome your difficulties? |  |
|  |  |  | ...been able to enjoy your normal day-to-day activities? |  |
|  |  |  | ...been able to face up to your problems? |  |
|  |  |  | ...been feeling unhappy or depressed? |  |
|  |  |  | ...been losing confidence in yourself? |  |
|  |  |  | ...been thinking of yourself as a worthless person? |  |
|  |  |  | ...been feeling reasonably happy, all things considered? |  |
| TEMPO Q1 | ASR | Among the descriptions below, indicate how they have applied to you in the past 7 days by ticking one of the boxes. If some of the descriptions do not apply to you, answer by thinking of similar situations you have encountered. | I’m too forgetful | 0 = Not at all true  1 = Sometimes or a little true  2 = Very true or often true |
|  |  |  | I have trouble concentrating or paying attention for long |  |
|  |  |  | I can’t get my mind off of certain thoughts |  |
|  |  |  | I have trouble sitting still |  |
|  |  |  | I cry a lot |  |
|  |  |  | I deliberately try to hurt or kill myself |  |
|  |  |  | I worry about my future |  |
|  |  |  | I don't eat as well as I should |  |
|  |  |  | I'm afraid of certain animals (dogs, insects, etc.) or certain situations (elevators, heights, crowds, etc.) |  |
|  |  |  | I feel worthless or inferior |  |
|  |  |  | I accidentally get hurt a lot, accident-prone |  |
|  |  |  | I hear sounds or voices that other people think  aren’t there |  |
|  |  |  | I’m impulsive or act without thinking |  |
|  |  |  | I’m nervous or tense |  |
|  |  |  | Parts of my body twitch or make nervous  movements |  |
|  |  |  | I feel too guilty |  |
|  |  |  | I feel tired without good reason |  |
|  |  |  | My heart is pounding or racing with no known medical cause |  |
|  |  |  | I fail to finish things I should do |  |
|  |  |  | There is very little that I enjoy |  |
|  |  |  | My work/school performance is poor |  |
|  |  |  | I would rather be with older people than  with people of my own age |  |
|  |  |  | I repeat certain acts over and over |  |
|  |  |  | I hear sounds or voices that other people think  aren’t there |  |
|  |  |  | I worry about my family |  |
|  |  |  | I sleep more than most other people  during day and/or night |  |
|  |  |  | I have trouble making decisions |  |
|  |  |  | I do things that other people think are strange |  |
|  |  |  | I have thoughts that other people would think are strange |  |
|  |  |  | I rush into things without considering  the risks |  |
|  |  |  | I think about killing myself |  |
|  |  |  | I have trouble sleeping |  |
|  |  |  | I don't have much energy |  |
|  |  |  | I’m unhappy, sad, or depressed |  |
|  |  |  | People think I’m disorganised |  |
|  |  |  | I feel that I can’t succeed |  |
|  |  |  | I tend to lose things |  |
|  |  |  | I feel restless or fidgety |  |
|  |  |  | I’m too impatient |  |
|  |  |  | I’m not good at details |  |
|  |  |  | In this moment, I feel relaxed |  |
| TEMPO Q2-Q9 | ASR | Among the descriptions below, indicate how they have applied to you in the past 7 days by ticking one of the boxes. If some of the descriptions do not apply to you, answer by thinking of similar situations you have encountered. | I cry a lot | 0 = Not at all true  1 = Sometimes or a little true  2 = Very true or often true |
|  |  |  | I worry about my future |  |
|  |  |  | I don't eat as well as I should |  |
|  |  |  | I feel worthless or inferior |  |
|  |  |  | I’m impulsive or act without thinking |  |
|  |  |  | I am nervous or tense |  |
|  |  |  | Parts of my body twitch or make nervous  movements |  |
|  |  |  | I feel too guilty |  |
|  |  |  | I feel tired without good reason |  |
|  |  |  | My heart is pounding or racing with no known medical cause |  |
|  |  |  | I worry about my family |  |
|  |  |  | I think about killing myself |  |
|  |  |  | I have trouble sleeping |  |
|  |  |  | I'm unhappy, sad or depressed |  |
|  |  |  | I feel that I can't succeed |  |
|  |  |  | I feel restless or fidgety |  |
|  |  |  | I'm too impatient |  |
|  |  |  | At the moment I'm relaxed |  |
|  |  |  | I’m too forgetful |  |
|  |  |  | I'm afraid of certain animals (dogs, insects, etc.) |  |
|  |  |  | I'm afraid of certain situations (elevators, heights, crowds, etc.) |  |
|  |  |  | I’m afraid of doing wrong |  |
|  |  |  | I feel that no one loves me |  |
|  |  |  | I have seizures |  |
|  |  |  | I feel that others are out to get me |  |
|  |  |  | I lack self-confidence |  |
|  |  |  | I worry about my relations with the others |  |
|  |  |  | I worry about my social relations with the opposite sex |  |
| MIND COVID | PHQ8-ADS | Over the past 2 weeks, how often have you been bothered by any of the following problems? | Little interest or pleasure in doing things? | 0 = Not at all  1 = Several days  2 = More than half the days  3 = Nearly every day |
|  |  |  | Feeling down, depressed, or hopeless? |  |
|  |  |  | Trouble falling or staying asleep, or sleeping too much? |  |
|  |  |  | Feeling tired or having little energy? |  |
|  |  |  | Poor appetite or overeating? |  |
|  |  |  | Feeling bad about yourself—or that you are a failure or have let yourself or your family down? |  |
|  |  |  | Trouble concentrating on things, such as reading the newspaper or watching television? |  |
|  |  |  | Moving or speaking so slowly that other people could have noticed? Or the opposite—being so fidgety or restless that you have been moving around a lot more than usual? |  |
|  |  |  | Feeling nervous, anxious, or on edge |  |
|  |  |  | Not being able to stop or control worrying |  |
|  |  |  | Worrying too much about different things |  |
|  |  |  | Trouble relaxing |  |
|  |  |  | Being so restless that it is hard to sit still |  |
|  |  |  | Becoming easily annoyed or irritable |  |
|  |  |  | Feeling afraid, as if something awful might happen |  |

Supplementary Figure 2: Flow chart of COMET, COVID and I, Mind COVID, and TEMPO samples, March 2020 - August 2022, n=13,635.

*The study's longitudinal design allowed for the collection of data from participants at various intervals, providing, for some, multiple observations for analysis.*
